# A Multi-hospital Study in Wuhan, China: Protective Effects of Non-menopause and Female Hormones on SARS-CoV-2 infection

**DOI:** 10.1101/2020.03.26.20043943

**Authors:** Ting Ding, Jinjin Zhang, Tian Wang, Pengfei Cui, Zhe Chen, Jingjing Jiang, Su Zhou, Jun Dai, Bo Wang, Suzhen Yuan, Wenqing Ma, Lingwei Ma, Yueguang Rong, Jiang Chang, Xiaoping Miao, Xiangyi Ma, Shixuan Wang

## Abstract

**Importance:** How to explain the better prognosis of female coronavirus disease 2019 (COVID-19) patients than that of males?

**Objective:** To determine the correlation between menstruation status/sex hormones and prognosis of COVID-19, and to identify potential protective factors for female patients.

**Design, Setting, and Participants:** A cross-sectional study of COVID-19 patients who were hospitalized at Tongji and Mobile Cabin Hospitals from Jan 28, 2020 to March 8, 2020.

**Exposures:** Confirmed SARS-CoV-2 infection.

**Main Outcomes and Measures:** Sex differences in severity and composite endpoints (admission to intensive care unit (ICU), use of mechanical ventilation, or death) of COVID-19 patients were compared. The correlation analysis and cox/logistic regression modeling of menstruation status/sex hormones and prognosis were conducted. Correlation between cytokines related to immunity and inflammation and disease severity or estradiol (E2) was revealed.

**Results:** Chi square test indicated significant differences in distribution of composite endpoints (p<0.01) and disease severity (p=0.05) between male and female patients (n=1902). 435 female COVID-19 patients with menstruation records were recruited. By the end of Mar 8, 111 patients recovered and discharged (25.3%). Multivariate Cox regression model adjusted for age and severity indicated that post-menopausal patients show the greater risk of hospitalization time than non-menopausal patients (relative hazard [RH], 1.91; 95% confidence interval [CI], 1.06-3.46) Logistic regression model showed that higher anti-müllerian hormone (AMH) as a control for age increases the risk of severity of COVID-19 (HR=0.146,95%CI = (0.026-0.824) *p=0.029*). E2 showed protective effect against disease severity (HR= 0.335, 95%CI = (0.105-1.070), p= 0.046). In the Mann-Whitney U test, the higher levels of IL6 and IL8 were found in severe group (*p= 0.040, 0.033*). The higher levels of IL2R, IL6, IL8 and IL10 were also observed in patients with composite end points (*p<0.001, <0.001, 0.009, 0.040*). E2 levels were negatively correlated with IL2R, IL6, IL8 and TNFα in luteal phase (Pearson Correlation=−0.592, −0.558, −0.545, −0.623; *p=0.033, 0.048, 0.054, 0.023*) and with C3 in follicular phase (Pearson Correlation=-0.651; *p=0.030*).

**Conclusions and Relevance:** Menopause is an independent risk factor for COVID-19. E2 and AMH are negatively correlated with COVID-19’s severity probably due to their regulation of cytokines related to immunity and inflammation.

**Key Points:** *Question:* Any differences in the outcomes between hospitalized female and male COVID-19 patients? If so, why?

*Findings:* Female patients display better prognosis than male patients. Non-menopausal women have shorter length of hospital stays, and AMH and E2 are negatively correlated with COVID-19’s severity. There is a negative correlation between E2 and the levels of IL6, IL8, IL2R and TNF-α, which are significantly correlated with disease severity or composite endpoint.

*Meaning:* Non-menopause and female sex hormones, especially E2 and AMH, are potential protective factors for females COVID-19 patients. E2 supplements could be potentially used for COVID-19 patients.

## Introduction

Starting from December 2019, COVID-19 has rapidly spread across the world, causing widespread concern. It’s worth noting that there was a definite sex difference in the morbidity and mortality of COVID-19. According to the recent data from the Chinese Center for Disease Control and Prevention (CDC), the ratio of male infection to female infection reached 2.7:1 ^1^. Among 72314 patients in China, the morbidity and case-fatality rates (CFR) (48.6% and 1.7%) of women were significantly lower than those of men (51.4%, 2.8%). More deaths were found in males than females (63.8% over 36.2%) ^2^. The mortality rate of women aged 15-49, under the average menopausal age 50, was even lower. These results indicate that females are less susceptible than males, and have better outcomes.

SARS-COV and MERS, “sisters” of SAR-CoV-2, also showed sex preference in their infection of humans with males experiencing higher CFR compared to females. ACE2 has been shown to be the receptor of both SARS-CoV and SARS-CoV-2. Interestingly, it is expressed more in male lungs than female lungs ^3^. Consistently, the expression of ACE2 is downregulated by estrogen E2 ^4^. This was confirmed in animal experiments, showing that estrogen mitigates the susceptibility and the severity of phenotype for SARS-CoV, while ovariectomy or intervention with estrogen receptor inhibitors increased the mortality of female mice ^5^. An anti-virus drug study showed that estrogen receptor inhibitors possess a great potency in antiviral activity screening against MERS-CoV, SARS-CoV and other RNA virus, and they also have an anti-Ebola virus effect in mice ^6^. These studies suggest that E2 might play a protective role in SARS-CoV or MERS-CoV infection with lower susceptibility, severity and better outcomes of their corresponding diseases. However, the effect of sex hormones and its related menstruation status on the progression and outcomes of SARS-CoV-2-caused COVID-19 remains unknown.

In our attempt to address this question, we enrolled COVID-19 patients at Tongji Hospital and Mobile Cabin Hospital. As described below, we found that there are the sex differences in the severity and outcomes of this disease. Also, we showed there is a good correlation between menstruation status/hormone levels and the severity or outcomes of COVID-19. Finally, by examining the association between cytokines related to immunity and inflammation and disease severity, clinical outcomes or E2, we learned that E2 is inversely correlated with the levels of these cytokines. Our results demonstrate that the better clinical outcomes of female COVID-19 patients are well correlated with menstruation status and female hormone levels.

## Methods

### Study design and participants

This collaborative clinical study included patients from three branches of Tongji Hospital (Sino-French New City Branch and Optical Valley Branch) and Mobile Cabin Hospital between Jan 28 and March 8, 2020. Tongji Hospital was one of the first batch of hospitals in Wuhan to receive severe and critically ill patients. The Mobile Cabin Hospital was mainly used to treat mild/moderate patients. All COVID-19 patients (n=1902) from the three branches were included to explore the sex differences. 509 female patients (441 of Tongji Hospital and 68 of Cabin Hospital) were inquired by telephone follow-up with menstrual status and gynecologic history, 435 patients were finally included. The correlation between menstruation status and disease severity or outcomes were analyzed. We also tested serum cytokines related to immunity and inflammation for 263 females and sex hormones levels for 78 female patients except those who denied our request. The flowchart of the study is shown in Figure 1.

**Figure 1.**
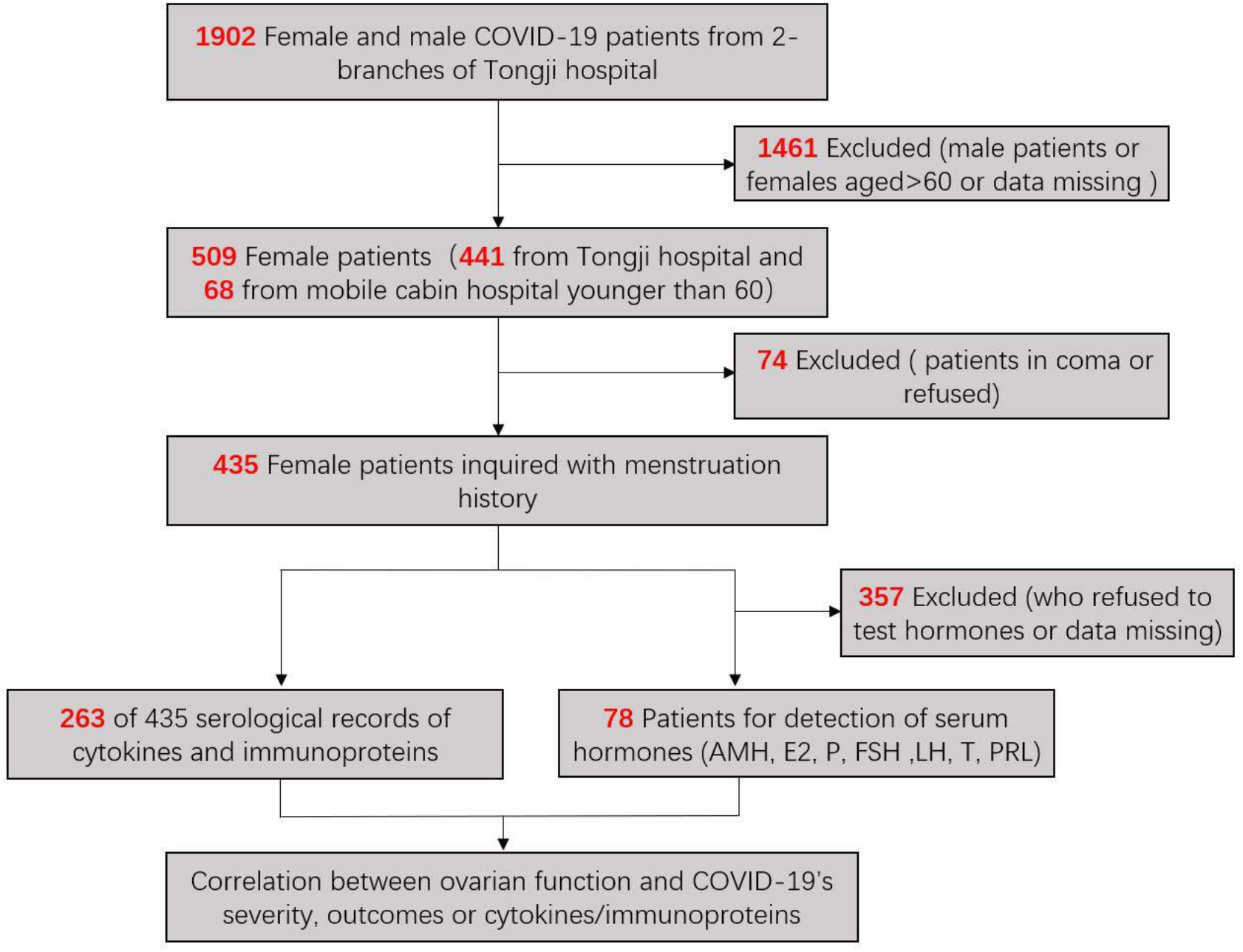
Flow chart of patient recruitment procedure. Patients (n=1902) from the three branches of Tongji Hospital were included to explore the sex differences in COVID-19’s prognosis. 509 female patients (441 of Tongji Hospital and 68 of Cabin Hospital) were inquired by telephone follow-up with menstrual status and gynecologic history. 435 patients with complete medical history were finally included. Then, we tested serum cytokines related to immunity and inflammation for 263 females and sex hormones levels for 78 female patients except those who denied our request. Finally, we determined the correlation between menstruation status/sex hormones and prognosis of COVID-19.

### Data Collection

COVID-19 was diagnosed based on the WHO interim guidance. A confirmed case of COVID-19 was defined as a positive result on real-time reverse-transcriptase– polymerase-chain-reaction (RT-PCR) assay of throat swab specimens^7^. Information recorded included exposure history, clinical symptoms, comorbidities, laboratory findings, chest computed tomographic (CT) scans and clinical outcome data. All laboratory testing was performed according to the clinical care needs of the patient. Laboratory tests included a complete blood count, C-reactive protein, procalcitonin, erythrocyte sedimentation rate, ferritin, immunoproteins including immunoglobulin A (IgA), immunoglobulin G (IgG), immunoglobulin M (IgM), complement 3 (C3) and C4, cytokines including IL-1β, IL-2R, IL-6, IL-8, IL-10 and TNF-α. The clinical outcomes (i.e., discharge, mortality, admission to ICU, mechanical ventilation, or death) were monitored up to March 8, 2020.

### Hormones Detection

Serum follicle-stimulating hormone (FSH), luteinizing hormone (LH), estradiol (E2), testosterone (T), prolactin (PRL), and progesterone (P) levels were measured using a chemoluminescence-based immunometric assay on an UniCelDxI 800 immunoassay system (Beckman Coulter, Inc, California, USA). Serum concentrations of AMH was measured using the Elecsys AMH kit (Roche, Inc, Basel, Switzerl). All these samples were measured in the same laboratory. The intra- and inter-assay coefficients of variation were all <15%. The lowest amount of AMH that could be detected with a 95% probability in a sample was 0.01 ng/mL.

### Ethics and Trial Registration

This study was reviewed and approved by the Medical Ethical Committee of Tongji Hospital of Huazhong University of Science and Technology (TJ-IRB20200214). Oral informed consent was obtained from each enrolled patient. The trial has been registered in Chinese Clinical Trial Registry (ChiCTR2000030015).

### Statistical analysis

Continuous variables were expressed as the means and standard deviations or medians and interquartile ranges (IQR) as appropriate. Categorical variables were summarized as the counts and percentages in each category. Mann-Whitney U test were applied to continuous variables, chi-square tests and Fisher’s exact tests were used for categorical variables as appropriate. Multivariate Cox proportional hazards models were used to evaluate the independent effect of menstruation regularity on prolonging hospitalization time, controlling age, severity and comorbidity. Multiple logistic regression model was used to assess the independent adjusted relationship between menstruation regularity and disease severity controlling age and E2 level. Pearson’s correlation analysis was used to explore the relationship between E2 and cytokines related to immunity and inflammation. All analyses were conducted with SPSS software version 19.0 and R software version 3.5. *P*<0.05 was considered significant. Data were analyzed by statisticians.

## Results

### Sex Difference in Severity and Composite Endpoint of COVID-19 Patients

To explore the sex difference in severity and outcomes, a cohort of patients (n=1902) infected with SARS-CoV-2 were recruited from the three branches of Tongji Hospital. There were more female patients in mild group (male 412, 44.06%; female 444, 45.92%), approximately the same proportion of female and male patients in severe group (male 486, 51.98%; female 503, 52.02%), and less female patients in critical group (male 37, 3.96%; female 20, 2.07%, p= 0.05). By the end of Mar 8, more female patients (97, 10.03%) than male patients (85, 9.09%)) had been discharged, and less female patients (16, 1.65%) than male patients (45, 4.81%) had died (p< 0.01). There was a significant difference between male and female patients in the severity and clinical outcomes, while not in the age distribution (eTable 1, eFigure 1). These results indicate that female patients are relatively milder and have better outcomes, which are in accordance with recent published articles ^1,2^.

### Characteristics of Female Patients Followed-up for Menstruation Status

The 509 patients from Tongji hospital and Mobile Cabin hospital were classified into non-severe group (386, 75.8%) and severe group (123, 24.2%) according to the American Thoracic Society guideline for community-acquired pneumonia ^8^ at admission and the published article of COVID-19 by Nanshan Zhong and his colleagues ^9^. The median age was 49 years (IQR, 38-56), female patients tended to be younger in non-severe group (median age 47 years) than in severe group (median age 52 years) (*p<0.001*) and more were suffered in 41-55y group (*p=0.022*). 52 patients had a body mass index more than 24, 138 patients stated their recent mental disorder, and 443 of 509 lived in Wuhan, there was no significant difference in disease severity or composite end point. At admission, most common symptoms were fever (393, 77.2%), cough (156, 30.6%) and dyspnea (54, 10.6%). 23.5% (119/506) patients had one or more comorbidities, 24.4% (90/250) patients had one or more gynecological diseases. 47.6% (206/433) patients received antiviral treatment, and 45.7% (198/433) patients received antibiotic treatment. By the end of Mar 8, 111 patients had been discharged, and 22 patients reached the composite endpoint (Table 1).

### The Relationship between Menstruation Status and COVID-19’s Severity or Outcomes

To determine the correlation between menstruation and disease progression, 435 patients who offered their complete menstruation history were included. In the last 6-12 months, 148 (34%) patients had regular menstrual cycle (21-35days), 36 (8.3%) patients had irregular menstrual cycle (shorter than 21days or longer than 35 days), and 251 (57.7%) patients had entered menopause (amenorrhea more than one year).

In non-severe group, patients with different menstruation status were 36.1%, 9.9% and 54.0% for regular, irregular and menopause, respectively. In the severe group, while the proportion of regular and irregular decreased to 27% and 3%, respectively, the proportion of menopause increased to 70% (*p=0.008*; eTable 2). But if age was considered as an adjust variable, the significance was reduced (*p= 0.533* for irregular group, *p= 0.079* for menopause group; eTable 2), even when the regular and irregular groups were merge into one non-menopause group (*p=0.260*; eTable 2). However, when discharge was used as an endpoint and variables including comorbidities, disease severity, age, and menstruation (non-menopause or menopause) were brought into a multivariate Cox regression analysis, menstruation showed definite protective effect. Patients with non-menopause tended to have lower hospitalization proportion as time passing by and to be discharged earlier than patients with menopause, control age and severity (relative hazard [RH]= 1.91; 95% confidence interval [CI], 1.06 – 3.46); eTable 2, Figure 2).

**Figure2.**
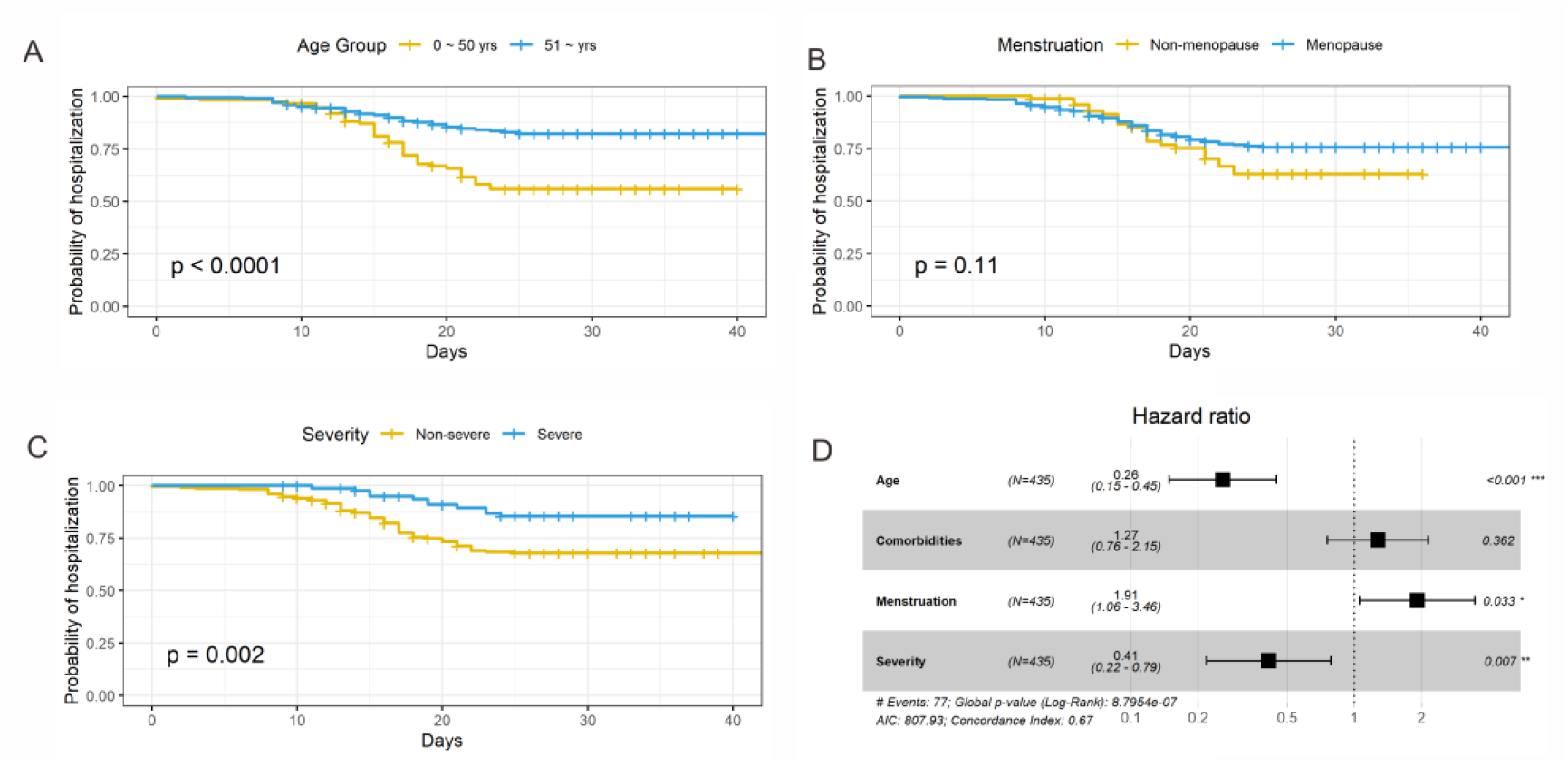
Cox analysis of Age, Menstruation Status and Disease Severity with Probability of Hospitalization. A, Univariate cox regression of age with probability of hospitalization; B, Univariate cox regression of menstruation status with probability of hospitalization, menstruation status was divided into non-menopause (regular or irregular) and menopause. C, Univariate cox regression of disease severity with probability of hospitalization. D, Multivariate Cox analysis, the covariates age, menstruation and severity are significant (*p < 0.001, 0.033, 0.007*). However, the covariate comorbidities fail to be significant (*p = 0.362*, which is greater than 0.05). The hazard ratio for menstruation is 1.91 indicating a strong relationship between non-menopauseand decreased risk of days in hospital. The hazard ratio for age and severityis 0.26 and 0.41 respectively, which indicate age and severity have a huge impact on risk of days in hospital.

**Figure3.**
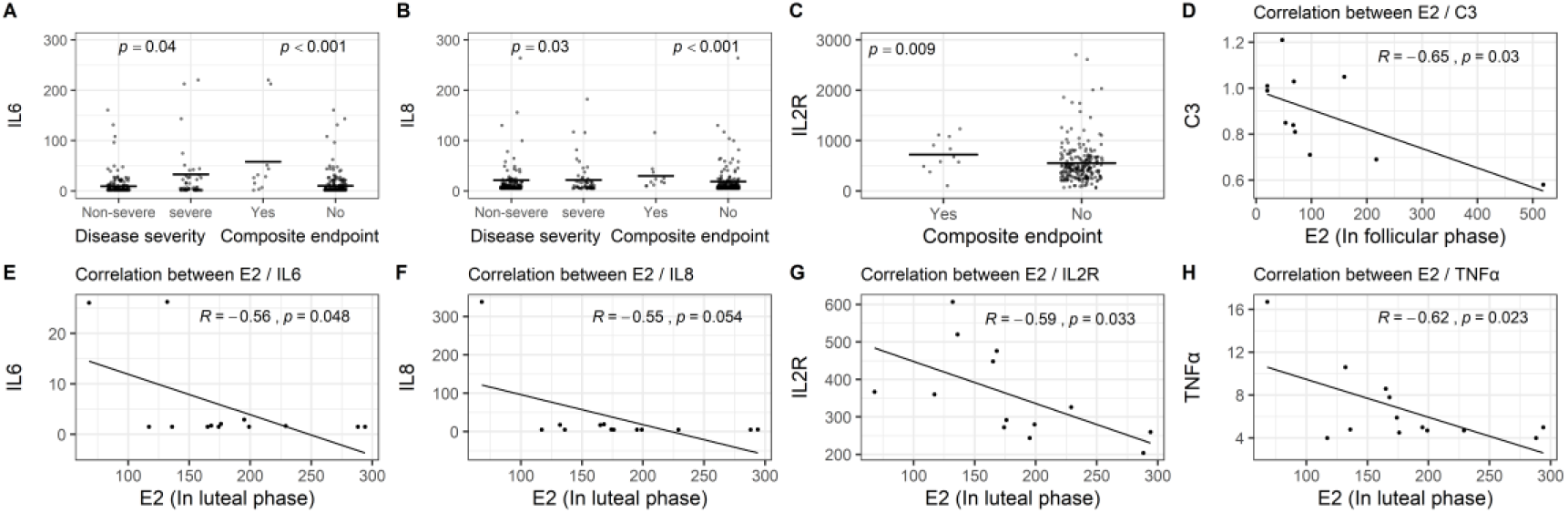
Correlation between Immunity/Inflammation related Cytokines and Disease Severity, Composite Endpoint and E2 in different phase. A, Correlation between IL6 with disease severity and composite endpoint. B, Correlation between IL8 with disease severity and composite endpoint. C, Correlation between IL2R with composite endpoint. In A, B and C, mean value is marked as solid line. Results of Mann Whitney u test indicate that patients in severe group showed higher level of IL6 and IL8 (p=0.04,0.03), and patients reached composite endpoint presents a higher level of IL6, IL8 and IL2R (p<0.001, <0.001,0.009). D, Correlation between C3 and E2 of COVID-19 patients in follicular phase (n=11). Result of Pearson’s correlation analysis indicates a significant inverse correlation between the E2 and C3 (r =0.65, p=0.030). E, Correlation between IL6 and E2 of COVID-19 patients in luteal phase (n=13). F, Correlation between IL8and E2 of COVID-19 patients in luteal phase (n=13). G, Correlation between IL2R and E2 of COVID-19 patients in luteal phase (n=13). H, Correlation between TNF-α and E2 of COVID-19 patients in luteal phase (n=13). Results of Pearson’s correlation analysis indicate a significant inverse correlation between the E2 level and IL6 (r=0.56, p=0.048), IL8 (r=0.55, p=0.054), IL2R (r=0.59, p=0.033) and TNF α (r=0.62, p=0.023).

### The Relationship between Serum Hormone Levels and COVID-19’s Severity

The baseline clinical characteristics of 78 patients agreed to test the serum hormone were shown in eTable 3. Age, phase of menstrual cycle, menstrual regularity in last 6 months and menstrual volumes in last 3 months were similar in terms of COVID-19’s severity and composite end points. The correlation between sex hormones and disease severity were shown in Table 2. E2, AMH, LH, T levels seemed to be most closely related to disease severity. Composite endpoints were not discussed for this group, since all patients were alive, not undergoing any of the composite endpoints at the end of observation. Since age affects sex hormones levels strongly, it was used as an adjust variable. In such, AMH still showed protective effect against disease severity (AHR= 0.146, *p= 0.029*), while LH and T did not (AHR= 2.388, *p=0.149*; AHR= 0.556, *p= 0.303*). E2 got a critical p value (AHR= 0.335, *p= 0.065*). Considering menstruation phase affected E2 levels greatly, it was used as another adjust variable with age. E2 showed a clear protective effect against the disease (AHR= 0.3, *p= 0.046*; Table 2).

### Correlation Analysis between Cytokines related to Immunity and Inflammation and E2 or COVID-19’ Severity/Outcomes

The correlation between cytokines related to immunity and inflammation and disease severity or composite end point were analyzed and shown in eTable 4. Higher levels of IL6 and IL8 were found in the severe group than that in non-severe group (*p= 0.040, 0.033;* Figure 5, eTable 4). Higher levels of IL2R, IL6, IL8 and IL10 were also observed in patients with composite end points than patients without composite end points (*p<0.001, <0.001, 0.009, 0.040;* eTable 5).

No significant correlation between the AMH level and these cytokines were found in this study. The correlation between the E2 level (either in luteal or follicular phase) and the level of these cytokines were shown in Figure 5 and Table S5. E2 levels were negatively correlated with that of IL2R, IL6, IL8 and TNFα in luteal phase (Pearson Correlation= −0.592, −0.558, −0.545, −0.623; *p= 0.033, 0.048, 0.054, 0.023*) and with C3 in follicular phase (Pearson Correlation= −0.651; *p= 0.030*; Figure 5, eTable 5). Collectively, the levels of IL6 and IL8 were positively correlated with COVID-19’s severity and composite endpoints. The E2 level was negatively correlated to that of IL6, IL8, IL2R and TNFα in luteal phase, and C3 in follicular phase.

## Discussion

It has remained incompletely understood if sex might significantly impact the outcomes of COVID-19 patients. Here, our study indicated the proportion of severe patients is less in females than that of males. Additionally, the clinical outcomes of female patients were better than that of male patients, which is in accordance with recent studies ^1,2,9,10^. Apparently, sexual dimorphisms account for differences in severity and outcomes of infectious diseases between females and males.

Previous study has indicated that menstruation status and hormonal fluctuation may affect disease symptom severity^11^. Our study as presented here showed that menstrual cycles and volumes are associated with the clinical severity and outcomes. But if age was considered as an adjust variable, the significance was reduced, even when the regular and irregular groups were merged into one non-menopause group. Interestingly, postmenopausal patients have longer length of hospital stays than do non-menopausal females, when considering age and severity. Accordingly, psychiatric hospitalizations for women are significantly greater within 5 days of their last menstrual period^12^. Normal menstruation regularity may be an important protective factor for COVID-19 patients. However, menstruation is often affected by many factors, such as medication, environment and anxiety, aside from the periodic fluctuation of sex hormones.

Sex-specific disease outcomes following virus infections are attributed to sex-dependent production of steroid hormones, different copy numbers of immune response X-linked genes, and the presence of disease susceptibility genes in males and females ^13^. Our data showed that E2 and AMH are positively correlated with the disease severity. E2 is primarily produced in ovaries and considered as the most prevalent and potent form of circulating estrogen. Estrogens are thought to protect non-menopausal women from hepatitis C virus and other pathogens ^14,15^. Also, ovariectomized and estrogen receptor antagonist treated female mice were more susceptible to SARS-CoV, which may be due to the protective effects of estrogen receptor signaling in SARS-CoV infection ^13^. AMH changes slightly with menstrual cycle and drugs and serves as the optimal marker for ovarian reserve and function. AMH levels were positively associated with the number of oocytes retrieved, ovarian response, E2 and testosterone ^16^. Logistics regression analyses showed that the levels of E2 and AMH in non-severe group are higher than that of the severe group, which may play vital roles in the progression of COVID-19. In this study, non-severe females have higher levels of testosterone than do severe patients. Previous studies have shown that testosterone has suppressive effects on immune system, such as on cytokine production and lymphocyte proliferation ^17^. However, after adjusted by age, the significance became unobvious. In the disease progression of COVID-19, testosterone may work by suppressing immune storms. Our results also indicated that E2 has a stronger protective effect against the potential negative effects from testosterone.

Previous studies have suggested that high doses of E2 may inhibit the production of inflammatory cytokines (e.g., IL-1β, TNF-α), whereas stimulation with E2 at its physiological level enhances their production^18,19^. After viral infection, proinflammatory cytokines/chemokines secreted by macrophages induced various antiviral mechanisms. Several clinical studies reported that levels of proinflammatory cytokines/chemokines, including IL-6, IL-8, IFN-γ, MCP-1, and IP-10, are highly increased in patients with SARS, some of which were correlated with ARDS, respiratory syncytial virus infections and human infection by avian influenza viruses ^20,21^. In our study, the levels of IL6 and IL8 were negatively related to both disease severity and composite end points, and IL2R and IL10 were related to composite end points, consistent with a recent study ^22^.

E2 has also been shown to regulate host immune response ^17,23-25^. Our results indicated that E2 is negatively associated with C3 in follicular phase, and IL2R, IL6, IL8 or TNFα in luteal phase. Cytokines and chemokines play vital roles in immunity and immunopathology during virus infection. The effects of E2 on the immune system are mainly due to its concentration, distribution, and different expression of two main estrogen receptor-ERα and ERβ in various lymphoid cells. E2 was involved in both cell-mediated and humoral immune responses. E2 can also initiate the differentiation from monocytes into inflammatory dendritic cells by regulating the expression of cytokines and chemokines and promote greater internalization and antigen presentation to naive T cells^26^. Thus, E2 may be a protective factor for female COVID-19 patients by regulation of cytokines or immunoproteins such as IL2R, IL6, IL8, TNFα and C3. Female symptoms severity and outcomes may change during times of hormone fluctuation.

## Limitations

This study has several limitations. First, correlation analysis showed that E2 is negatively correlated with COVID-19’s severity, but the causal relationship between them remains unclear. Second, the serum E2 level was just from once blood sample regardless of phase of menstrual cycle, so that the results may not completely reflect the average E2 level and its effects. In order to alleviate this error, we divided menstrual cycle into follicular and luteal phase when analyzed with E2. Third, the sample size of serum sex hormones, 78, might not be large enough. More samples are needed to further determine the relationship of E2 level with the outcomes.

## Conclusions

In summary, our data revealed that sex-bias does exist in COVID-19 patients, and females are less severe and have better outcomes than do males, which is probably in some extent due to the protective role of non-menopause and sex hormones, especially E2, by regulation of cytokines related to immunity and inflammation. Our study provides a new perspective to understand the susceptibility, progression and prognosis of COVID-19’s sex difference. The levels of sex hormones and the status of menstruation can serve as potential markers for the clinical prognosis and outcomes of COVID-19. E2 supplement may have great potency in anti-virus infection, which need to be further studied in animal experiments and clinical researches.

## Data Availability

The raw/processed data required to reproduce these findings cannot be shared at this time as the data also forms part of an ongoing study.

## Author contributions

S.Wang and X.Ma had full access to all of the data in the study and take responsibility for the integrity of the data and the accuracy of the data analysis. The final version of the article to be published was approved by all authors. T Ding, J Zhang, T Wang, P Cui and Z Chen contributed equally and share first authorship. S.Wang and X.Ma share corresponding authorship.

Concept and design: S.Wang, X Ma, T Ding, J Zhang, T Wang, P Cui and Z Chen. Acquisition, analysis, or interpretation of data: T Wang, J Zhang, P Cui, Z Chen, J Jiang, S Zhou, S Yuan, W Ma, L Ma, J Jiang, B Wang, J Dai. Drafting of the manuscript: T Ding, J Zhang, T Wang, P Cui and Z Chen. Critical revision of the manuscript for important intellectual content: Y Rong, X.Ma and S.Wang.

Statistical analysis: J Chang, X Miao.

Supervision: X.Ma and S.Wang.

## Conflict of Interest Disclosures

No one reported.

## Funding/Support

This work was financially supported by the Clinical Research Pilot Project of Tongji hospital, Huazhong University of Science and Technology (No. 2019CR205).

## Role of the Funder/Sponsor

The funders had no role in the design and conduct of the study; collection, management, analysis, and interpretation of the data; preparation, review, or approval of the manuscript; and decision to submit the manuscript for publication.

## Notes

### Competing Interest Statement

The authors have declared no competing interest.

### Clinical Trial

ChiCTR2000030015

